# Minimal mRNA uptake and inflammatory response to COVID-19 mRNA vaccine exposure in human placental explants

**DOI:** 10.1101/2023.02.01.23285349

**Authors:** Veronica Gonzalez, Lin Li, Sirirak Buarpung, Mary Prahl, Joshua F. Robinson, Stephanie L. Gaw

**Author notes:** **Correspondence:** Stephanie L. Gaw, MD, PhD, Associate Professor, Division of Maternal-Fetal Medicine, Department of Obstetrics, Gynecology and Reproductive Sciences University of California, San Francisco, 513 Parnassus Ave, Box 0556 16HSE, San Francisco, CA 94143, Joshua F. Robinson, PhD, Assistant Professor, Center for Reproductive Sciences, Department of Obstetrics, Gynecology and Reproductive Sciences, University of California, San Francisco, 513 Parnassus Ave, Box 0556 16HSE, San Francisco, CA 94143. These authors contributed equally to this work (Veronica Gonzalez, Lin Li).

## Abstract

Despite universal recommendations for COVID-19 mRNA vaccination in pregnancy, uptake has been lower than desired. There have been limited studies of the direct impact of COVID-19 mRNA vaccine exposure in human placental tissue. Using a primary human villous explant model, we investigated the uptake of two common mRNA vaccines (BNT162b2 Pfizer-BioNTech or mRNA-1273 Moderna), and whether exposure altered villous cytokine responses. Explants derived from second or third trimester chorionic villi were incubated with vaccines at supraphysiologic concentrations and analyzed at two time points. We observed minimal uptake of mRNA vaccines in placental explants by in situ hybridization and quantitative RT-PCR. No specific or global cytokine response was elicited by either of the mRNA vaccines in multiplexed immunoassays. Our results suggest that the human placenta does not readily absorb the COVID-19 mRNA vaccines nor generate a significant inflammatory response after exposure.

## Introduction

Despite strong recommendations for vaccination against COVID-19 in pregnancy by all major public health agencies and professional societies ^1,2^, vaccination rates have been low in this vulnerable population ^3^. As pregnant individuals were initially excluded from clinical trials ^4,5^, and there is a lack of studies examining the direct impact of COVID-19 mRNA vaccines on the placenta.

The placenta acts as a semi-protective barrier against certain pathogens, environmental chemicals, and pollutants; however, many compounds, small-molecules, viral agents, etc., are known to cross the placenta to varying degrees ^6^. COVID-19 mRNA vaccines deliver the mRNA encoding SARS-CoV-2 spike protein encapsulated by lipid nanoparticles ^7^. While cellular uptake is necessary to induce systemic immunity, whether this lipid-mRNA complex can transfer across the placental barrier remains unknown.

Even in the absence of transplacental transmission, exposures may stimulate placental inflammatory responses. The human placenta has the capacity to express all known cytokines, thought to be mainly produced by immunogenic fetal cell populations, *e*.*g*., fetal macrophages (Hofbauer cells), syncytiotrophoblasts and cytotrophoblasts ^8^. However, in general, pregnancy is a tolerogenic condition and the placental immune environment tends to be shifted towards an anti vs. pro-inflammatory state. Aberrant inflammatory responses to drugs or microorganisms have been associated with adverse pregnancy outcomes such as fetal growth restriction, preterm birth, and preeclampsia ^9^. Imbalances in cytokine levels may also negatively impact fetal brain development, increasing the risk of long-term neurodevelopmental disorders such as hyperactivity, autism spectrum disorders and schizophrenia ^10-13^.

The exogenous mRNA vaccine is immunostimulatory by design and can be recognized by a variety of cell surface receptors and endosomal sensors such as Toll-like receptors 7 (TLR7) and TLR8, resulting in potent immune activation and production of type I interferons and other inflammatory cytokines and chemokines ^7,14^. Intradermal injection of the mRNA vaccine produces both local and systemic inflammation ^15^, as evidenced by mild to moderate pain at the injection site as well as fever, headache and chills within 7 days after injection. Endogenous placental responses to direct mRNA vaccine exposures are unknown.

In this study, we investigated the capacity of mRNA vaccine uptake in human placenta tissue after direct exposures *in vitro* using a primary human chorionic explant model in comparison with common human placental/non-placental cell lines. Additionally, using the chorionic explant model, we evaluated whether the presence of the mRNA vaccine alters release of cytokines important in placental inflammatory response.

## Results

### mRNA vaccine uptake after *in vitro* exposures in human placental explants and trophoblast cell lines

Human explants from second and third trimester placentas and four human cell lines─two of placental origin─BeWo placental choriocarcinoma cells and JEG-3 choriocarcinoma cells, and two other cell lines, A549 lung carcinoma cells and 293T embryonic kidney cells, were incubated with either mRNA-1273 (Moderna) or BNT162b2 (Pfizer) vaccines at two concentrations (0.1 µg/mL or 1 µg/mL) or durations (0.5 and 4h). Total RNA was isolated from explant tissue and/or cell pellets and analyzed by RT-PCR against vaccine-specific Spike mRNA sequence as previously described ^16^. Standard curves were generated with known concentrations of the two mRNA vaccines, mRNA-1273 and BNT162b2, and cycle threshold (CT) values corresponding to > 0.05 pg/µL, the estimated lower limit of detection for both vaccines, were reported as positively detected (mRNA-1237; CT < 23.2; BNT162b2; CT < 24.6; **Figure 1A**). In general, vaccine mRNA was minimally or not detected in villous explants exposed to either of the mRNA vaccines with either 0.1 µg/mL or 1 µg/mL exposures. In contrast, both vaccines were detected in BeWo, JEG-3, 392T and A549 cell lines at varying amounts in a dose- and time-dependent manner (**Figure 1B**). For example, in JEG-3 exposed to 0.1 µg/mL of mRNA-1273, we did not detect the mRNA vaccine at 0.5h (mean < 0.05 pg/mL), however, by 4h it was found at 0.223 pg/uL. In JEG-3 exposed to 1 µg/mL, we detected a mean concentration of 0.37 pg/µL at 0.5h, and these levels increased to 2.116 pg/µL at 4h. In general, higher levels (average 3.3-fold) of mRNA-1273 were detected in all four cell lines than those exposed to BNT162b2 at equivalent concentrations and exposure durations. Exceptions were noted for BeWo (1 µg/mL BNT162B2) and A549 (1 µg/mL mRNA-1273), in which detectable vaccine levels at 4h versus 0.5h. In general, our results suggest that there is a concentration and time (i.e., exposure duration) dependence on the amount of vaccine mRNA detected.

**Figure 1.**
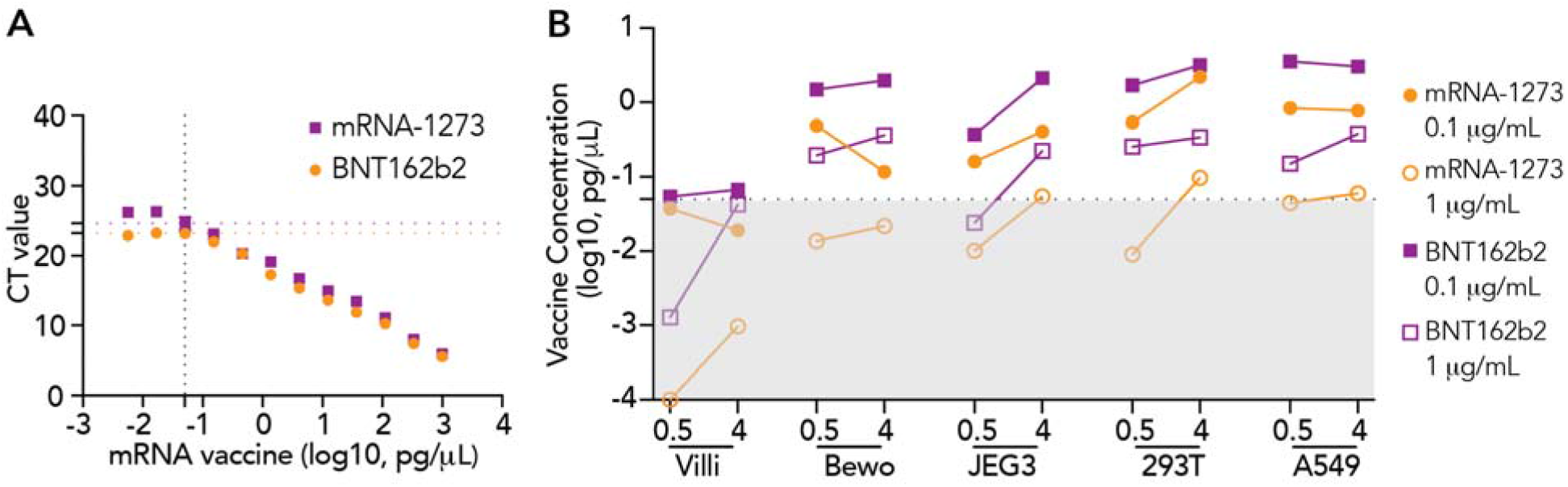
Detection of vaccine-derived Spike protein mRNA in human placental explants and four human cell lines using qRT-PCR. (A) Vaccine messenger RNA (mRNA) standard curves. Standard curves for mRNA-1273 and BNT162b2 vaccines were generated to calculate vaccine-derived Spike mRNA concentrations in placental explants and cells. The limit of detection (LOD) level was set according to the standard curve (0.05 pg/µL, mRNA-1237 CT = 23.2, BNT162b2 CT = 24.6). (B) Vaccine mRNA detected in placental explants (villi) and four different types of cell lines exposed to mRNA-1273 or BNT162B2 at 0.1 µg/mL or 1 µg/mL for 0.5h or 4h. Results were calculated by extrapolating CT values based on the derived standard curves for each mRNA vaccine. Black dotted line represents the LOD level of 0.05 pg/µL. Grey box represents the area under LOD. All data represent 2 independent experiments. CT = Cycle threshold.

### Detection of mRNA COVID-19 vaccines by RNAscope in situ hybridization (ISH) in placental explants

To further analyze the degree of uptake and tissue localization of the mRNA vaccines into placental explants, we used RNAscope-based in situ hybridization assay─a method proposed to be more sensitive than qRT-PCR ^17^ ─to detect the vaccine-related mRNA. Explants cultured from second trimester or term placentas were incubated with mRNA-1273 or BNT162b2 vaccines, at 0.1 µg/mL or 1 µg/mL for 0.5 or 4h, respectively. Explants cultured in optimized medium without mRNA vaccines were used as a control group. Peptidylprolyl isomerase B (PPIB, positive control) and DapB (negative control) probes were used to interpret the results (**Figure 2A** and **2B**). At 0.5 or 4h, in explants exposed to either of the two vaccines, we did not find a positive signal corresponding to the detection of either mRNA-1273 or BNT162b2 in the placental explants (n=2; **Figure 2C)**.

**Figure 2.**
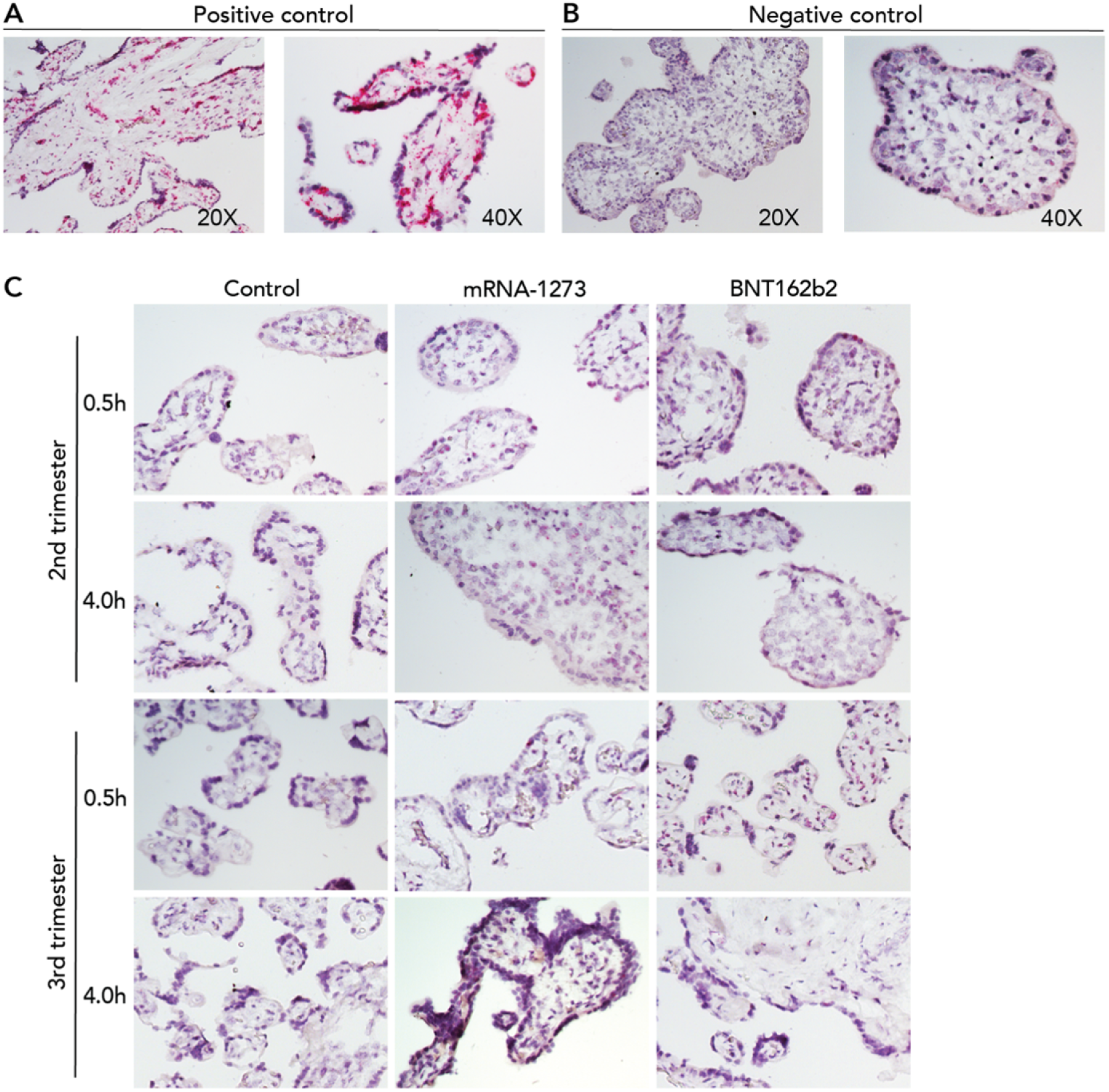
Lack of detection of COVID mRNA vaccine via in situ hybridization in placental explants. Chorionic villi explants derived from 2nd (n=2) or 3rd trimester (n=2) human placentas were incubated with 0.1 µg/mL (not shown) or 1 µg/mL mRNA-1237 or BNT162B2 vaccines. After 0.5h or 4h, tissues were fixed, paraffin-embedded, sectioned and probed for mRNA vaccine using RNAscope in situ hybridization. (A) Positive and (B) negative controls for RNAscope detection of mRNA vaccine. Peptidylprolyl isomerase B (PPIB) was used as a positive control. Pink dots corresponding to PPIB mRNA can be observed within the chorionic villi at 20X and 40X. DapB was used as a negative control. No signal was detected at 20 or 40X. (C) In situ detection of mRNA vaccine in vaccine-exposed explants. No signal was evident in explants incubated with either vaccine at any of the two timepoints.

### mRNA vaccines stimulation of cytokine expression in placental explants

Even in the absence of significant uptake into placental tissue, exogenous exposures may induce cytokine secretion that could have an adverse impact on the placenta/fetus ^18^. Thus, we utilized a Luminex assay to quantify 24 unique cytokines in the conditioned media of second trimester and term explants treated with either of the two mRNA vaccines at 0.1 µg/mL or 1 µg/mL for 0.5 or 4h. Standards for each cytokine were used to interpolate concentrations in the conditioned media. Of the 24 cytokines tested, four (IL-1β, IL-17A, IL-12p70, IFN-γ) were not detected in any samples in the interpolable range. Levels of different cytokines varied widely; IL-7 had the lowest level (median 2.48 pg/mL) while TNF-R1 had the highest level (median 13258.41 pg/mL) (**Figure 3A**). Levels of cytokines (IL-5, IL-28A, IL-28B, CXCL9, CCL17 and CCL22) with low variance (< 15%) were excluded from downstream analyses to reduce detection of false positives.

**Figure 3.**
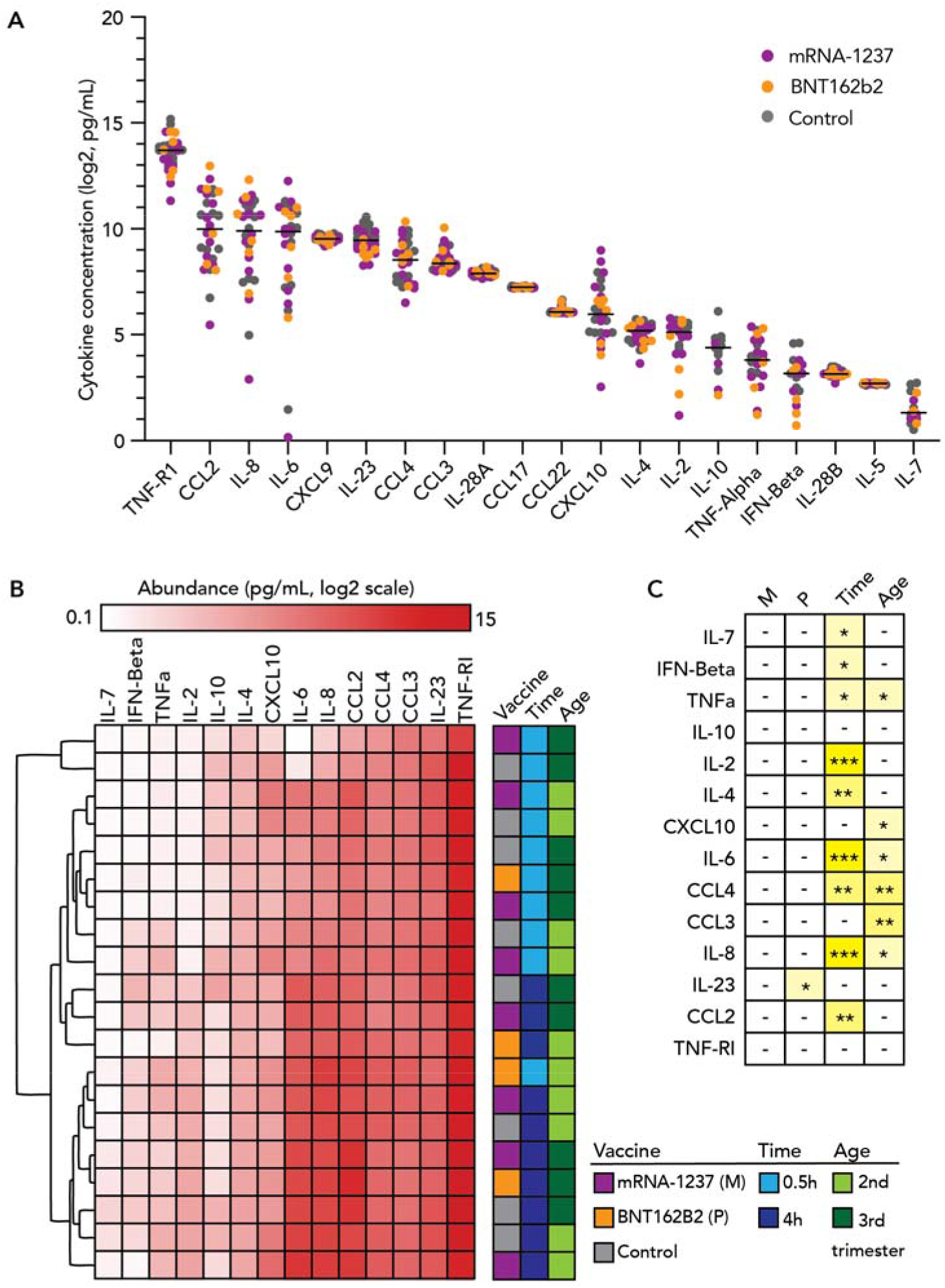
Expression of inflammatory mediators in conditioned media of placental explants incubated with COVID mRNA vaccine. Chorionic villi explants derived from 2nd or 3rd trimester human placenta (n=6) were incubated with 0.1 µg/mL or 1 µg/mL mRNA-1237 (n=4) or BNT162B2 (n=2) vaccines for 0.5h or 4h. Conditioned media was collected and levels of 20 unique cytokines were measured using the Luminex multiplex assay. (A) Distribution in abundance of cytokines in all samples. Black bars represent median levels in cytokine abundance. (B) Hierarchical clustering of samples by levels of cytokines. We excluded cytokines of low variance from this analysis (n=6 cytokines). In general, sample profiles clustered by culture time (0.5 vs. 4h). (C) Significance of factors in cytokine expression evaluated using multiple linear regression models. Asterisks represent significance cutoffs: p<0.05 (*); p<0.005 (**); and p<0.0005 (***). Abbreviations listed for factors assessed in our analysis: mRNA-1237 (M), BNT162B2 (P) vaccines, culture time (Time), or trimester (Age).

We assessed the remaining 14 cytokines in the context of vaccine exposure, exposure duration or gestational age of the explants. We applied unsupervised hierarchical clustering analysis to determine if global patterns in cytokine profiles segregated by one of the three variables (**Figure 3B**). Notably, the majority of samples clustered by exposure duration, indicating that this factor was most influential in affecting cytokine levels. In general, patterns in cytokine levels were not associated with either of the two vaccines in comparison with the control or gestational age.

In the next step of our analysis, we implemented multiple linear regression models to assess whether specific cytokines were associated with exposure to one of the two vaccines while adjusting for culture duration. In general, significant changes in cytokine levels were not observed with mRNA-1273 exposure (**Figure 3C**). These observations were similar with BNT162b2 exposure, where overall we did not see cytokine induction to be associated with vaccine exposure vs. control, with the exception of IL-23 which was modestly decreased at the two time points (∼ -0.2 pg/mL; p=0.03. **Supplemental Figure 1**). Of note, several cytokines were associated with culture duration or gestational age irrespective of vaccine exposure. IL-2, IL-4, IL-6, IL-8, CCL2 and CCL4) increased in abundance in the conditioned media over culture time. Specific cytokines seem to be released higher into the culture medium in 2^nd^ trimester vs. term explants such as IL-8, CCL3, CCL4, TNF-α, CXCL10 and IL-6 (**Figure 3C**). In general, vaccine exposures did not considerably modify cytokine abundance in vaccine-exposed vs. control groups.

## Discussion

To our knowledge, this is the first study to directly investigate the effects of COVID-19 mRNA vaccines in a human villous explant model of the placental barrier. We report minimal uptake of two commonly used COVID-19 mRNA vaccines. Additionally, direct exposure of explants to COVID-19 mRNA vaccines did not profoundly affect cytokine release, suggesting that human placental chorionic villi are resistant to mRNA vaccine penetration and interactions that lead to changes in inflammatory status.

We implemented two approaches to detect mRNA vaccine uptake: qRT-PCR and in situ hybridization. Following a 30min or 4h duration, vaccine mRNA was detected in monolayer trophoblast cell lines BeWo and JEG-3, which lack the cell-to-cell interactions of different cell types in tissue, and also may not perfectly recapitulate the syncytiotrophoblast barrier (**Figure 1C**). In contrast, there was very limited (or none at all) detectable vaccine mRNA in the placental explants. As a secondary measure, we applied RNAscope in situ hybridization which provides additional information regarding the localization of the mRNA vaccine within the tissue architecture of explants. Using this method, we did not observe vaccine mRNA in the chorionic villi, including syncytiotrophoblast, intravillous cytotrophoblasts or other cells that comprise the villous stroma. This suggests that the placental architecture and inherent cellular relationships serve as an effective barrier, potentially blocking transfer of circulating mRNA vaccine across the placental syncytium. The syncytiotrophoblast layer is considered a robust physical and immunologic barrier that prevents hematogenous transmission of both virus and nonviral pathogens ^19-21^. In our study, we did not detect either mRNA vaccine in the syncytiotrophoblast layer (after 0.5 or 4h of exposure). The lipid nanoparticle delivery system is designed to facilitate membrane fusion and intracellular delivery of the vaccine ^22,23^. It is unknown how the placental tissue resists lipid nanoparticle uptake, despite direct exposure of fresh vaccine in the culture media. Different lipid nanoparticle composition has been recently shown to impact the degree of placental uptake ^24^. We would anticipate that *in vivo*, the uptake of mRNA vaccines into the placenta would be less due to many other factors that likely contribute to vaccine degradation. In summary, our study suggests limited uptake of the mRNA vaccine in chorionic villi.

Several cytokines were identified to increase in the conditioned media in a time-dependent manner irrespective of vaccine treatment or gestational age (**Figure 3C**). Therefore, we implemented a statistical approach to account for temporal differences to determine if specific cytokines were evoked by the mRNA vaccines in placental explants compared to the no-vaccine control group. In general, the presence of either mRNA vaccine did not lead to significant perturbations in the release of cytokines into the media. These results suggest that the mRNA vaccines are not significantly immunogenic to the placenta. Thus, we provide evidence that the fetus is unlikely to experience a placental or fetal immune response to the vaccine itself which may allay persistent concerns about vaccine effects on the fetus. This is in contrast to COVID-19 infection, which is known to alter placental and fetal immune status, even in mild infections ^25^.

The human placenta is complex and unique in its anatomy and function, as such, there are limitations in the ability of animal models to replicate human placentation and pregnancy ^26^. Human chorionic explants models maintain the natural structure and cellular organization of chorionic tissue that cell lines lack, permitting the investigation of tissue responses *in vitro* ^27^. In this study, we incubated tissue explants with the vaccine for up to several hours, with the goal of exaggerating the impact of vaccines on the chorionic villus and associated cell populations. We chose selected supraphysiologic concentrations (0.1 or 1 µg/mL) based on estimated local circulating levels of mRNA vaccines in mice vaccinated for influenza, which reported maximum concentrations of mRNA vaccines detected in the spleen and liver to be 0.087 µg/mL and 0.047 µg/mL, respectively ^28^. Local concentrations at the maternal-fetal placental barrier are likely to be lower than the testing concentrations used in this study. Under physiological conditions, vaccine concentrations are low in peripheral blood following tissue absorption, hematologic circulation, and natural degradation.

Another advantage of our human explant model is the ability to examine the tissue responses at different gestational ages, as the placenta matures to support the different developmental stages of the fetus. Different patterns of cytokine secretion were observed between placenta from different trimesters (**Figure 3C**). For example, there were generally higher levels of IL-8, CCL3, CCL4, TNF-α, CXCL10 and IL-6 secreted from second trimester placental explants compared with those from third trimester. However, future studies may address this question with a larger sample size due to the potential dynamics in sensitivity to inflammatory stimuli across pregnancy.

Our results provide further support to published clinical research that COVID-19 vaccination during pregnancy was not associated with increased pregnancy complications or adverse neonatal outcomes ^29,30^. There have been no adverse histopathologic findings in placentas isolated from vaccinated pregnancies ^31,32^. Thus, our results provide additional reassurance regarding the safety of COVID-19 mRNA vaccines in pregnancy, which have been shown to protect both the mother and the baby against COVID-19 and associated complications ^33-37^.

## Data Availability

All data produced in the present work are contained in the manuscript.

## Acknowledgements

We thank Kenneth Scott (University of California, San Francisco, UCSF Health Pharmacy) and Hannah J. Jang, PhD, RN, PHN, CNL (University of California, San Francisco, UCSF School of Nursing and the Center for Nursing Excellence and Innovation) for providing unused vaccine for this study.

## Author Contributions

VG, JR and SG conceived and designed the study. VG, LL, SB, MP and JR conducted the experiments. LL, MP, JR and SG analyzed the data. VG and LL wrote the first draft of the manuscript. JR and SG supervised the experiments. All authors provided critical editing of the manuscript.

## Methods

### Cell Culture

BeWo placental choriocarcinoma cells were cultured in F-12K medium (Gibco, Waltham, MA). JEG-3 choriocarcinoma cells, A549 lung carcinoma cells and 293T embryonic kidney cells were cultured in Dulbecco’s modified Eagle’s medium (DMEM; Gibco). All media were supplemented with 10% fetal bovine serum and 1% penicillin-streptomycin. Twenty-four hours prior to exposure to mRNA vaccines, cells were seeded at a density of 3 × 10^5^ cells per well in a six-well plate and cultured at 37°C and 5% CO_2_ in a humidified incubator. On the next day, after discarding the old media and washing the cells with DPBS, complete culture media with BNT162b2 or mRNA-1273 at the concentration of 0.1 µg/mL or 1 µg/mL were separately added to each well. After incubation (30 minutes or 4 hours), media was removed and cells were washed with DPBS twice and then lysed in RTL (Qiagen, Hilden, Germany).

### Tissue specimens

The study was approved by the institutional review board of the University of California, San Francisco. Second trimester placentas without evidence of pathology were obtained from consenting participants following termination of pregnancy. Third trimester placentas from uncomplicated pregnancies were obtained following late preterm or term (36-39 weeks gestational age) planned cesarean deliveries. Indications for early delivery included history of classical cesarean section, history of uterine surgery, or placenta previa. Written consent was obtained from all participants. Placental biopsies measuring 3 × 3 cm were collected, washed in Cytowash (see below) and stored on ice prior to proceeding with explant dissection.

### Placental explant cultures

Placental biopsies were washed in Cytowash (consisting of DME/H-21 (Gibco), 2.5% fetal bovine serum (Hyclone), 1% glutamine plus (Atlanta Biologicals), 1% penicillin/streptomycin (Invitrogen), and 1% gentamicin (Gibco) ^38^ and dissected into 1.0 cm^2^ chorionic villi explants. A single villus was placed in one well of a 24-well plate containing 0.5 ml of prewarmed cell free media [DME/H-21, 2% Nutridoma (Roche), 1% sodium pyruvate (Sigma), 1% Hepes buffer (Invitrogen), 1% glutamate plus (Atlanta Biologicals), and 1% penicillin/streptomycin (Invitrogen)] with 10% fetal bovine serum and incubated at 37°C in 5% CO2/95% for 30 minutes. Vaccine mixtures for explant exposure were then prepared such that final vaccine concentrations were: BNT162b2 and mRNA-1273 vaccine 0.1 µg/mL and 1 µg/mL, as well as control medium. Testing concentrations were based on estimates of localized circulating levels of mRNA vaccines in organs of mice vaccinated for influenza ^28^. Explants were exposed for 0.5 or 4 hours and incubated at 37°, after which 200 ul of supernatant were collected and stored at -4°C for cytokine analysis. Exposed explants were washed in PBS and then stored at -80 °C until RNA extraction, or formalin-fixed for 24 hours at room temperature, then washed in 70% ethanol and stored at - 4°C until undergoing paraffin embedding.

### RNA extraction and real-time PCR (RT-PCR)

Total RNA was extracted from cells and villi using the RNeasy Micro Kit (Qiagen) RNA quantity and quality were measured by a NanoDrop spectrophotometer. Complementary DNA (cDNA) synthesis was performed using qScript cDNA Synthesis Kit (Quantabio, Beverly, MA), followed by qPCR using SYBR™ Green qPCR Master Mix with QuantaStudio 6 Flex system (Applied Biosystems, Waltham, MA). Each sample was run in duplicates. Primer sequences for both BNT162b2 and mRNA-1273 were as follows: AACGCCACCAACGTGGTCATC (forward, 5’ to 3’) and GTTGTTGGCGCTGCTGTACAC (reverse, 5’ to 3’). Vaccine concentrations in the cells and villi were determined by interpolation on the standard curve made from BNT162b2 and mRNA-1273 vaccines as previously described ^16^. Mann-Whitney test to determine statistical significance between vaccine-treated groups and control groups. P values below 0.05 are considered to be statistically significant.

### RNAscope

The antisense probe targeting the spike region (21,631-23,303) of SARS-CoV-2 Wuhan-Hu-1 strain (GenBank accession number NC_045512.2) were purchased (Advanced Cell Diagnostics [ACD], Newark, CA). Sequence alignment of the probe and the mRNA vaccine sequence showed 73-79% identity. RNAscope^®^ 2.5 Assay was preformed according to the manufacturer’s protocol. Briefly, following formalin-fixed paraffin-embedded sample preparation and pretreatment, gene-specific probe pairs were hybridized to target mRNA. Probe targeting the spike region were hybridized to a cascade of signal amplification molecules, culminating in binding of HRP-or AP-labeled probes. Fast Red substrate was added to detect target RNA in tissue. Visualization of target RNA was preformed using standard bright field microscope. Peptidylprolyl isomerase B (PPIB, positive control) and DapB (negative control) probes were used as the positive and negative controls.

### Cytokine assay

Concentrations of 24 cytokines, IFN-γ, IFN-β, TNF-α, IL-1β, IL-10, IL-17A, IL-2, IL-4, IL-5, IL-6, IL-7, CXCL8/IL-8, IL12p70, IL-23, TNF-RI, CCL17/TARC, CCL2/MCP-1, CCL22/MDC, CCL3/MIP-1α, CCL4/MIP-1b, CXCL9/MIG, CXCL10/IP-10, IL-28A or IL-28B, were measured using the custom human premixed multiplex magnetic Luminex assay (Bio-Techne, Minneapolis, MN) according to the manufacturer’s protocol. Briefly, supernatant from placental explant cultures treated with vaccines were mixed with microparticle cocktail and added to each well of a 96-well plate. Plate was incubated at room temperature on a shaker at 800 rpm for 2 hours, followed by 1h incubation with biotin-antibody and 30-minutes incubation with streptavidin-PE. Plate was washed with wash buffer 3 times after each incubation. Then wash buffer was add to wells for resuspending the microparticle and plate was read on a Luminex MAGPIX CCD Imager. All samples were run in duplicate, and standards and blanks were applied to each plate. Median fluorescence intensity (MFI) of each cytokine was corrected by subtracting background and interpolated to the standard curve. We applied unsupervised hierarchical clustering of cytokine levels using average linkage and Euclidean distance to determine whether global patterns associated with culture duration, vaccine exposure, or gestational age ^39^. Using GraphPad Prism 9.1.0 (GraphPad, San Diego, CA), we implemented multiple regression models to determine the significance of these variables on levels of specific cytokines: 1) BNT162b2 and culture duration; 2) mRNA-1273 and culture duration; or 3) culture duration and gestational age. P-values <0.05 were interpreted as significant.

**Supplemental Figure 1.**
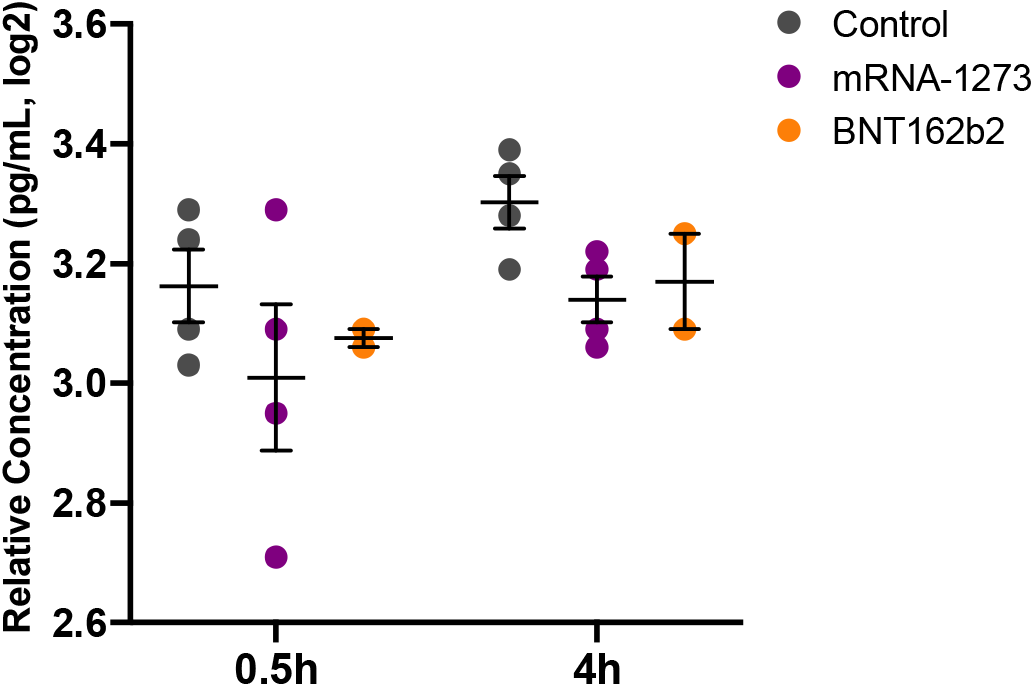
Expression of IL-23 in conditioned media of placental explants incubated with COVID mRNA vaccines.

